# Ageing and Dementia: Age-Period-Cohort Effects of Policy Intervention in England, 2006-2016

**DOI:** 10.1101/2021.03.13.21253508

**Authors:** Kamila Kolpashnikova

**Affiliations:** University of Oxford, 42 43 Park End St., Oxford, OX2 6UD United Kingdom

**Keywords:** dementia, National Dementia Policy, Dementia Challenge, policy evaluation

## Abstract

**Background:** Dementia is one of the most important challenges of our time. According to the Dementia Statistics Hub, only about 66% of all UK residents with dementia were diagnosed in 2017-2018. Yet, there are reservations about the early diagnosis of dementia-related diseases. As a result, the UK National Screening Committee does not recommend systematic population screening of dementia.

**Methods:** This study added additional evidence of the effectiveness of the National Dementia Strategy and increased numbers of diagnosis of dementia on the younger cohorts of the elderly, using the intrinsic estimator age-period-cohort models and the English Longitudinal Study of Ageing data.

**Results:** Age effects show that screening and diagnosis increases in volume only among those aged 75 and above, suggesting that many of the younger elderly were not diagnosed. Period effects show that although there was an initial increase due to new policy implementation, the trend stalled in later years, indicating that the increase might not have been even across the period when controlled for age and cohort. The study also shows that cohort effects indicate lower prevalence in younger cohorts controlled for age and period.

**Conclusions:** Although more research in diverse contexts is warranted, this study cautions against the abandonment of timely diagnosis and increased screening and shows some effectiveness of prevention strategies on the national level.

## Background

According to Dementia Statistics Hub, one in three people with dementia are left undiagnosed in the UK [1]. Between 2009 and 2015, the UK put tremendous efforts in addressing dementia in domestic health policy, including increasing diagnosis rates, promoting awareness and prevention, and changing treatment strategies [2]. This study attempts to contribute to the previous research and evaluate UK health policy effects on dementia diagnosis using the age-period-cohort analysis. It is particularly important to evaluate the effects of the national policy and strategy change, given the urge to evaluate its adequacy was expressed in recent work [3, 4].

The National Dementia Strategy (NDS) was launched in February 2009 to improve awareness about the illness, encourage an increase in diagnosis, prevention, and the quality of care for people diagnosed with dementia-related diseases. In 2012, NDS was superseded by Dementia Challenge, which was then updated to Dementia 2020 Challenge in 2015.

Among other changes with the UK-wide dementia policy, more people with mild dementia were prescribed acetylcholinesterase inhibitors, whereas antipsychotic prescriptions continued to decrease [2]. K Donegan, N Fox, N Black, G Livingston, S Banerjee and A Burns [2] report that the prescription of antidementia medication doubled in percentage in between 2005 and 2015 and reached 36.3%, whereas the antipsychotic drug prescription halved from 22.1% to 11.4%. Research shows that acetylcholinesterase inhibitors may slow down the cognitive decline in patients with Alzheimer’s disease, though more research is warranted [5, 6].

Although the present paper does not aim to call for the reversal of the current screening mandate in the UK, since the harmful effects of the early diagnosis need to be addressed and clear ethical and procedural measures outlined in the communication with the patients and caregivers [4], it aims to provide new investigative evidence whether any changes were observed due to the implementation of the dementia-related national policy, particularly NDS and Dementia Challenge of 2012-2015. This aim is achieved by using improved methods in age-period-cohort analysis, the intrinsic estimator (IE) models and the English Longitudinal Study of Ageing, 2006-2017. The IE models allow disentangling age, period, and cohort effects to illustrate whether the effects are constant or diverging over age groups, period, and cohorts.

As the age effects are expected to follow the national recommendations on screening, this study is mostly interested in the analysis of period and cohort effects. The invariable period effects would suggest that there were no screening and diagnosis increase in the period, controlled for age and cohort, which we know is not factual in the case of the diagnosis of dementia in the analysed period [1]. On the one hand, the invariant cohort effects would confirm the assumption that prevention strategies, as well as the concurrent changes in treatment strategies, had no effect on the younger cohorts of the elderly. On the other hand, changes in the cohort trends might indicate some evidence of the opposite.

## Methods

### Data Source

The English Longitudinal Study of Ageing (ELSA) is used for the analysis [7] and was accessed through the UK Data Services. ELSA survey collection started in 2002 and interviewed people aged 50 years and older. There were refreshment samples added to keep the study’s representativeness. The survey waves for ELSA were scheduled for every other year, but the actual collection spread over the two years. This project uses Wave 3 (2006) through 8 (2016) to include a few years prior to the introduction of the NDS in 2009 and a few years after the Dementia Challenge of 2012-2015. The total ELSA sample in the selected waves was 59,807 people. After restricting the sample to those aged 60 and above, the analytical sample included 42,848 (72% of the total sample) people.

### Outcome Variable: Prevalence of Dementia-related Disease

The dependent variable is measured by whether the diagnoses of dementia-related diseases, including Alzheimer’s disease, were reported at the surveyed wave. The outcome is a dummy variable, 1 for ‘yes’ and 0 for ‘no’.

Table 1 summarizes the dementia-related disease prevalence rates by age group and cohorts. It shows that the prevalence rate increases with age and that it is higher in older cohorts, as expected. The table also shows that the increased diagnosis in the pre-2015 period reported more cases of dementia for younger elderly (in between 65 and 74) and among those who were above 80 years of age.

**Table 1.** Prevalence Rates by Age and Period and Cohort and Period, in %

|  | Period |  |  | Total |
| --- | --- | --- | --- | --- |
|  | 2006-2009 | 2010-2014 | 2015-2017 |  |
| Dementia by Age Group (%) |  |  |  |  |
| 60-64 | 0.61 | 0.41 | 0.43 | 0.49 |
| 65-69 | 0.50 | 0.74 | 0.61 | 0.65 |
| 70-74 | 0.51 | 1.15 | 0.63 | 0.84 |
| 75-79 | 1.18 | 1.74 | 2.37 | 1.70 |
| 80+ | 1.83 | 4.75 | 3.79 | 3.65 |
| Dementia by Cohorts (%) |  |  |  |  |
| Before 1930 | 1.83 | 0 | 0 | 1.83 |
| 1930-1934 | 1.18 | 4.75 | 0 | 3.10 |
| 1935-1939 | 0.51 | 1.74 | 3.79 | 1.62 |
| 1940-1944 | 0.50 | 1.15 | 2.37 | 1.13 |
| 1945-1949 | 0.61 | 0.74 | 0.63 | 0.68 |
| 1950-1954 | 0 | 0.41 | 0.61 | 0.47 |
| 1955-1959 | 0 | 0 | 0.43 | 0.43 |
| Total | 13346 | 21304 | 8198 | 42848 |

### Age, Period, and Cohort

The analysis is based on the five-year intervals of the categories of age, period, and cohort, which are usually used in the analysis of APC effects [8–10]. The intervals of five years were used in the construction of age and cohort categories. However, the three resulting period categories were constrained by the actual years of the survey, 2006-2017. Thus, the resulting period categories were 2006-2009, 2010-2014, and 2015-2017.

### Analytical Strategy

This paper’s models employ the intrinsic estimator (IE) to disentangle age-period-cohort effects in dementia-related diseases [8, 11]. The IE modelling in age-period-cohort analysis remains the most appropriate way to analyse the effects without having to impose constraints on either age, period, or cohort categories [9, 10, 12]. Some critiques of the method exist. For instance, L Luo [13] showed that IE models would not work in all situations, using simulations. However, later, RK Masters, DA Powers, RA Hummer, A Beck, S-F Lin and BK Finch [12] showed that the situations where IE models will not work are very unlikely to happen in the real world and reclaimed the confidence in IE models in age-period-cohort analysis.

In public health research, Bell’s HAPC models, developed in A Bell [14], are used more commonly than the IE models. However, the HAPC models require strong assumptions regarding (usually) period effects [15]. Considering that period effects are expected to vary between 2006 and 2017 in England’s dementia prevalence trends, the use of IE models is preferred in the present paper.

## Results

Table 2 presents the results from the IE models on dementia-related diseases’ prevalence. Most of the age-period-cohort effects are significant in the model. Model 1 shows that the prevalence increases with age and period and decreases with a cohort change, except for the cohort born during WWII and the oldest cohort.

**Table 2.** Intrinsic Estimator Age-Period-Cohort Effects Models on Dementia-Related Diseases Prevalence

|  | Coefficients | SE | Confidence Interval |
| --- | --- | --- | --- |
| Age Group |  |  |  |
| 60-64 | -0.403*** | (0.119) | [-0.635, -0.170] |
| 65-69 | -0.535*** | (0.119) | [-0.767, -0.303] |
| 70-74 | -0.477*** | (0.121) | [-0.714, -0.239] |
| 75-79 | 0.286** | (0.093) | [0.103, 0.469] |
| 80+ | 1.128*** | (0.092) | [0.948, 1.309] |
| Period |  |  |  |
| 2006-2009 | -0.347*** | (0.078) | [-0.500, -0.194] |
| 2010-2014 | 0.172* | (0.067) | [0.040, 0.304] |
| 2015-2017 | 0.175* | (0.081) | [0.017, 0.333] |
| Cohorts |  |  |  |
| Before 1930 | -0.102 | (0.144) | [-0.384, 0.179] |
| 1930-1934 | 0.350** | (0.118) | [0.118, 0.581] |
| 1935-1939 | 0.157 | (0.119) | [-0.076, 0.390] |
| 1940-1944 | 0.458** | (0.143) | [0.177, 0.739] |
| 1945-1949 | 0.138 | (0.137) | [-0.130, 0.406] |
| 1950-1954 | -0.434** | (0.145) | [-0.717, -0.150] |
| 1955-1959 | -0.567 | (0.354) | [-1.261, 0.127] |
| Constant | -4.661*** | (0.080) | [-4.818, -4.504] |
| Observations | 42848 |  |  |
| Deviance | 5211.749 |  |  |
| Log-Lik. | -2605.875 |  |  |

Standard errors in parentheses. + $p < 0.10$ , \* $p < 0.05$ , \*\* $p < 0.01$ , \*\*\* $p < 0.001$

## Discussions

### Age Effects

Age effects based on the IE model in Table 3 are presented in Figure 1. The trends reveal that the diagnosis (preceded by screening) mostly happen among people of aged 75 and above.

**Figure 1.**
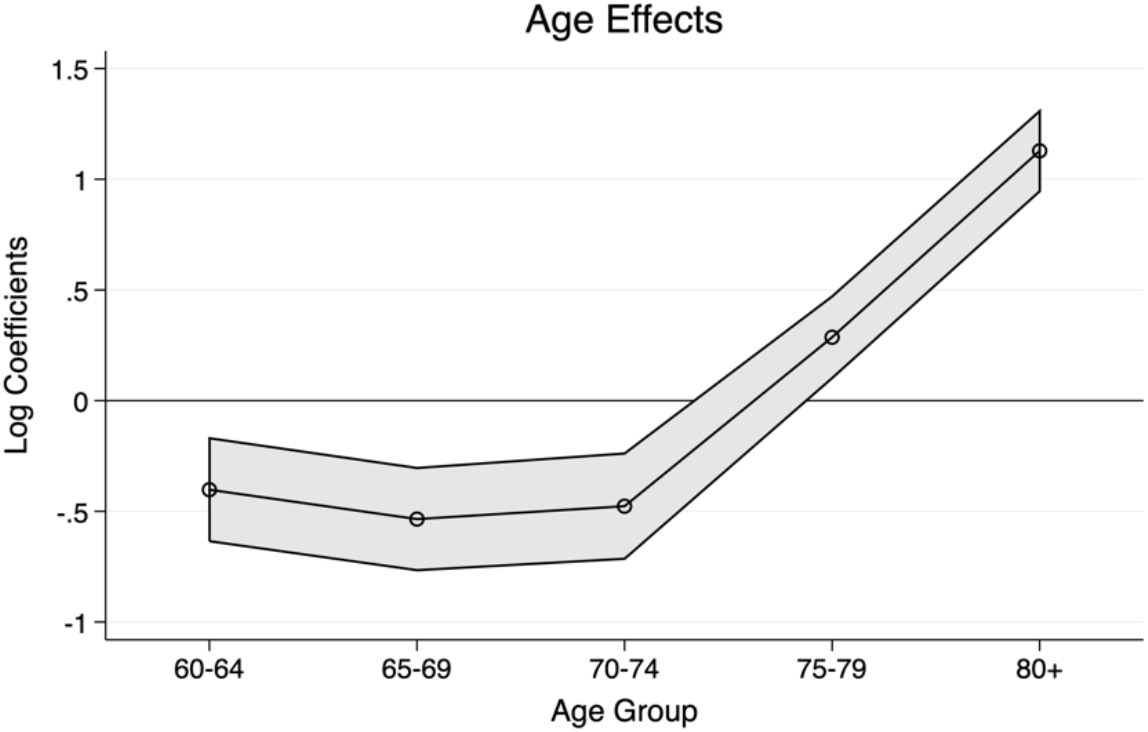
Age Effects.

This indicates that even during the analysed period since the start of the NDS with a policy mandate for increased testing of dementia, those aged 74 and below were rarely screened and diagnosed with dementia. The descriptive findings in Table 1 confirm this result for people of age 69 and below. For those between 70 and 74, the prevalence rate doubled between the first and the second period and reverted to the first period’s levels in the third period. These results suggest that very few younger elderly are diagnosed in a timely manner, even after the NDS, reflecting the current recommendations of the UK National Screening Committee. The steep increase for those aged 75 and above on the age effects confirms that many of these cases might remain undetected earlier if we assume that the prevalence increases gradually rather than exponentially over age groups. These results of the age effects are expected and are according to the national guidelines on the screening and diagnosis of dementia-related diseases.

### Period Effects

Period effects, presented in Figure 2, confirm that the period after 2009 since the inception of NDS was associated with increased dementia diagnosis (prevalence) in the population. The period effects, however, stalled since the scanning mandate remained only for the older elderly. The trends show a steep increase between the first period (2006-2009), followed by stagnation between 2010-2014 and 2015-2017 when controlled for age and cohort effects.

**Figure 2.**
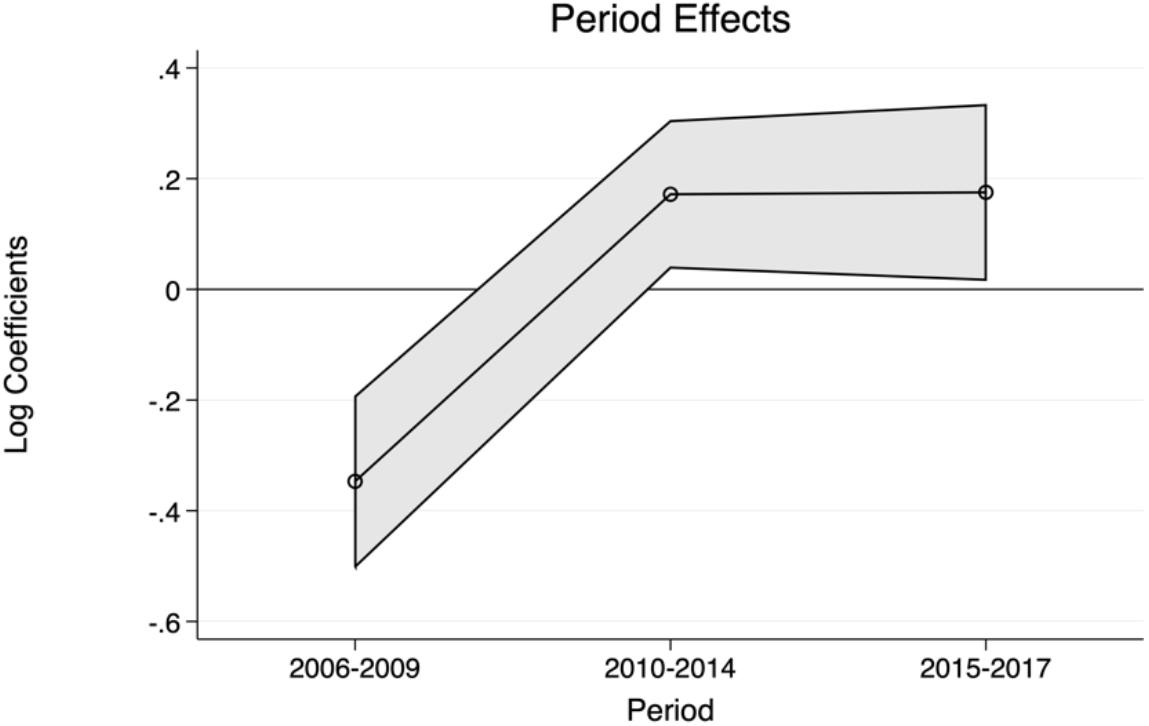
Period Effects.

**Figure 3.**
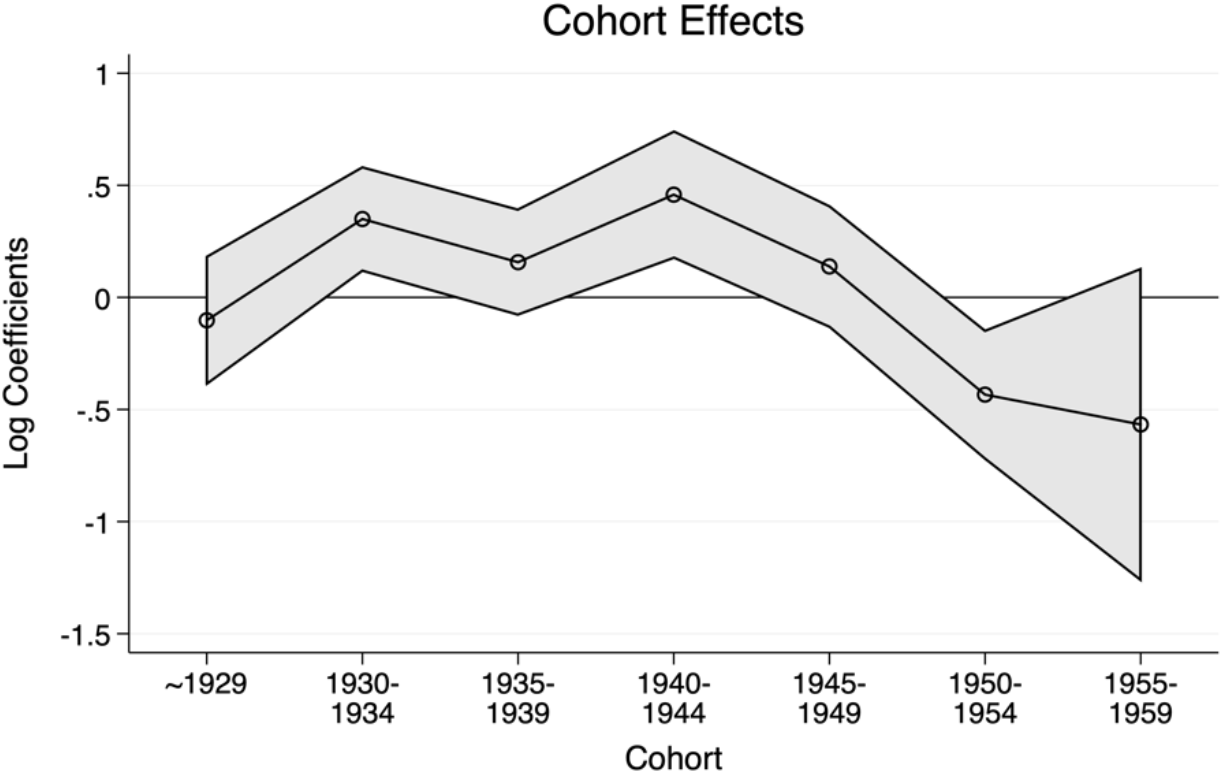
Cohort Effects.

Overall, the period effects confirm that the call for increased diagnosis in dementia was delivered since the implementation of the NDS but stalled in recent years.

### Cohort Effects

The cohort effects in this study can potentially show the effectiveness of the dementia risk prevention policy if a decreasing trend in prevalence is detected in earlier cohorts, net of age and period effects. The trends show that the risk prevention activities, treatment strategy changes, as well as changes in the lifestyle of the younger cohorts not related to the policy implementation might have positively affected the younger cohorts, particularly those born between 1950-1954. However, striking differences in prevalence can be confirmed between the younger cohort in the survey and those who are ten years older than them, which gives us a bit of optimism in terms of the effects of the risk prevention measures, although the association might be explained by other factors unrelated to the policy implementation as well. More extended observation is warranted to be able to say anything definitively.

## Conclusions

This study provides additional evidence of the effectiveness of the UK National Dementia Strategy and Dementia Challenge of 2012-2015, particularly of the timely diagnosis of dementia and risk prevention, based on the intrinsic estimator age-period-cohort analysis. The study provides a novel way to use APC models to analyse the effectiveness of preventive and diagnostic measures using longitudinal data. The findings confirm that age effects follow the screening recommendations installed on the national level. The period effects reveal a substantial increase in diagnosis following the implementation of the National Dementia Strategy and consequent stall in the trend in the later years. The cohort effects show some improvement in younger cohorts, suggesting the effectiveness of preventive policy or generational changes in lifestyles. However, the evidence presented in this study is not conclusive because of the limited data, and more research is warranted, particularly using a range of data from diverse contexts. The way to do this is to analyse the APC effects across different contexts, depending on their screening strategies and selected preventive measures, including in European, North American, Asian, and other countries.

One limitation of this study is that the prevalence of early detection of dementia-related diseases in the younger cohorts can also be related to other cohort-related changes: lifestyles and decreases in alcohol consumption and smoking, among other things.

Future research could focus on the effects of preventive measures and increased diagnosis over longer periods of time, particularly with the focus on mild cognitive impairment screening over time.

## Data Availability

ELSA data is accessible through UKDS

## Acknowledgements

Not applicable.

## Availability of data and materials

The English Longitudinal Study of Ageing is available through the UK Data Service.

## Declarations

### Ethics approval and consent to participate

Not applicable. This study is based on secondary data analyses of publicly available data.

### Consent for publication

Not applicable.

## References

1. Dementia Statistics Hub [https://www.dementiastatistics.org]

2. Donegan K, Fox N, Black N, Livingston G, Banerjee S, Burns A: Trends in diagnosis and treatment for people with dementia in the UK from 2005 to 2015: a longitudinal retrospective cohort study. The Lancet Public Health 2017, 2(3):e149–e156.

3. Brayne C, Kelly S: Against the stream: early diagnosis of dementia, is it so desirable? BJPsych bulletin 2019, 43(3):123–125.

4. Lohmeyer JL, Alpinar-Sencan Z, Schicktanz S: Attitudes towards prediction and early diagnosis of late-onset dementia: a comparison of tested persons and family caregivers. Aging & Mental Health 2020:1–12.

5. Giacconi R, Giuli C, Casoli T, Balietti M, Costarelli L, Provinciali M, Basso A, Piacenza F, Postacchini D, Galeazzi R: Acetylcholinesterase inhibitors in Alzheimer’s disease influence Zinc and Copper homeostasis. Journal of Trace Elements in Medicine and Biology 2019, 55:58–63.

6. Small DH: Acetylcholinesterase inhibitors for the treatment of dementia in Alzheimer’s disease: do we need new inhibitors? Expert opinion on emerging drugs 2005, 10(4):817–825.

7. Banks J, Batty GD, Coughlin K, Deepchand K, Marmot M, Nazroo J, Oldfield Z, Steel N, Steptoe, Wood M et al: English Longitudinal Study of Ageing: Waves 0-8, 1998-2017. In., 29th Edition edn: UK Data Service; 2019.

8. Yang Y, Schulhofer-Wohl S, Fu WJ, Land KC: The intrinsic estimator for age-period-cohort analysis: what it is and how to use it. Am J Sociol 2008, 113(6):1697–1736.

9. Schwadel P: Age, period, and cohort effects on religious activities and beliefs. Social Science Research 2011, 40(1):181–192.

10. Schwadel P, Stout M: Age, period and cohort effects on social capital. Soc Forces 2012, 91(1):233–252.

11. Yang Y, Fu WJ, Land KC: A methodological comparison of age-period-cohort models: the intrinsic estimator and conventional generalized linear models. Sociological methodology 2004, 34(1):75–110.

12. Masters RK, Powers DA, Hummer RA, Beck A, Lin S-F, Finch BK: Fitting age-period-cohort models using the Intrinsic Estimator: assumptions and misapplications.Demography 2016, 53(4):1253–1259.

13. Luo L: Assessing validity and application scope of the intrinsic estimator approach to the age-period-cohort problem. Demography 2013, 50(6):1945–1967.

14. Bell A: Life-course and cohort trajectories of mental health in the UK, 1991–2008–a multilevel age–period–cohort analysis. Social Science & Medicine 2014, 120:21–30.

15. Canizares M, Hogg-Johnson S, Gignac MA, Glazier RH, Badley EM: Increasing trajectories of multimorbidity over time: birth cohort differences and the role of changes in obesity and income. The Journals of Gerontology: Series B 2018, 73(7):1303–1314.

